# Validation of Registry-Based Indicators for Postdiagnostic Antibiotic Decisions in Pediatric Febrile Urinary Tract Infection

**DOI:** 10.64898/2026.03.19.26348369

**Authors:** Kalle Garpvall, Alex Aljundi, Anna Dahl, Ellinor Sterky, Joachim Luthander, Susanne Sütterlin

## Abstract

**Background:** Electronic prescribing registries are widely used for antimicrobial stewardship surveillance. Existing indicators predominantly measure structure or process, while validated outcome indicators remain rare. The present study evaluates how well rule-based measures capture clinically meaningful postdiagnostic antibiotic decision making in pediatric febrile urinary tract infection.

**Methods:** We conducted a retrospective, multicenter validation study including all empirically treated febrile UTI episodes across three Swedish pediatric emergency departments. Prescribing outcomes were classified using registry rules and compared with outcomes determined by clinician review and laboratory findings. Guidance Ratio (GR) and Discontinuation Ratio (DR) were calculated monthly and in aggregate for both clinically validated- and registry rule classifications.

**Results:** In total, 909 febrile UTI episodes were included across all sites. The rule-based GR was 49%. GR increased consistently with stronger diagnostic evidence. Among the 431 episodes with clinician-adjudicated follow-up, 63% resulted in guided treatment; 28% discontinued treatment, and 9% lacked follow-up documentation. The rule-based algorithm showed a sensitivity of 0.78 and a specificity of 1.00 for identifying guided outcomes. Monthly rule-based GR tracked validated temporal patterns but underestimated absolute values. A calibration function substantially improved agreement.

**Conclusions:** Rule-based indicators captured overall prescribing patterns but underestimated the level of prescribing concordant with guidelines. Validation against clinician reviewed reference data enabled calibration and improved the interpretability of indicators based on registry data for antimicrobial stewardship.

## Introduction

Antimicrobial stewardship has generated a wide range of quality metrics to monitor antibiotic use. The most applied indicators focus on prescription volumes, treatment duration, and the distribution of antibiotic classes, reflecting aspects of care that can be reliably derived from routine prescribing data. These indicators primarily capture antibiotic exposure as a result of prescribing decisions rather than the clinical reasoning that led to treatment initiation or continuation. Although developments in electronic health records have made it easier to access laboratory data, diagnostic codes and longitudinal follow-up; translating such information into robust and scalable metrics that focus on clinical decisions remains challenging. [1,2]

Febrile urinary tract infection (UTI) is one of the most common indications for empirical antibiotic treatment in pediatric emergency care. In young children, diagnosis is frequently uncertain because of nonspecific symptoms and limitations in obtaining representative urine samples. This frequently leads to initiation of antibiotics before microbiological confirmation. [3] Once urine culture results become available, clinicians are expected to reassess the diagnosis and decide whether empirical therapy should be continued, modified, or discontinued. This postdiagnostic decision point is central to antimicrobial stewardship but is rarely captured by existing prescribing indicators. [4]

Pediatric febrile UTI involves a relatively consistent sequence of clinical and microbiological information for diagnostic reassessment. This structure makes them particularly suitable for evaluating prescribing decisions using algorithms. However, classifications derived from registry rules require clinical validation to ensure that they reflect decisions based on clinical assessment. In this study, we aimed to validate registry rule classifications of prescribing outcomes for pediatric febrile UTI against outcomes determined by clinician review of medical records. Furthermore, the study aimed to assess whether ratio indicators can meaningfully quantify antibiotic decision making after reassessment.

## Material and Methods

### Setting

Antibiotic prescription data were evaluated from three Swedish pediatric emergency departments located at the Children’s Hospital of Uppsala University Hospital in Uppsala, and Karolinska University Hospital in Solna and Huddinge, Stockholm. All three pediatric emergency departments are university-affiliated tertiary care centers that manage most febrile pediatric UTIs within their respective catchment areas, including referrals from primary and secondary care.

Pediatric UTIs are managed according to current Swedish national guidelines. In brief, children presenting with suspected febrile UTI were tested with plasma C-reactive protein (CRP) and urine dipstick. Positive urine samples were sent for quantitative culture to the clinical microbiology departments. Empirical antibiotic treatment was initiated, and patients were reevaluated when culture results became available, typically within two to three days. The attending physician reassessed the patient (typically remote contact), interpreted the clinical and microbiological findings, and determined whether the diagnosis of febrile UTI was confirmed and if continued antibiotic treatment was warranted. Children aged 0 - 18 years were eligible for inclusion if empirical antibiotic treatment for presumed febrile UTI was initiated. Patients occasionally managed in outpatient care were not included.

According to Swedish regulations, informed consent was not required for this type of retrospective quality-register validation. The study was approved by the Swedish Ethical Review Board (document ID 2020-03297).

### Study design

*Data source and registry structure.* This was a retrospective, register-based validation study. Data were retrieved from the Swedish national antibiotic prescribing registry (“Infektionsverktyget”), which records the clinical indication and links each antibiotic prescription to the patient’s electronic medical record through the unique national personal identifier. The registry categorizes UTIs as febrile or afebrile. For cases registered as febrile UTI, data were extracted for both the initial empirical antibiotic treatment and any subsequent guided treatment prescribed for the same episode. The study period covered all consecutive visits during the period September 1, 2021 to August 31, 2023 for Uppsala: December 9, 2022 to May 31, 2023 for Solna, and December 9, 2022 to June 9, 2023 for Huddinge, respectively.

*Case identification and validation*. Eligible episodes were identified from Infektionsverktyget and all included visits underwent full electronic medical record review in the pediatric emergency departments to validate registry entries and extract clinical data. Clinical variables included body temperature (°C), plasma C-reactive protein (CRP, mg/L), and urine dipstick results. The latter included leukocyte esterase (0 to +4), erythrocytes (0 to +3), and nitrite (negative/positive). Urine culture results were reviewed for bacterial growth, species identification, and colony counts according to standard laboratory procedures. For the Uppsala dataset specifically, clinical follow-up documentation was available and used to establish a reference classification of prescribing outcomes. This was considered as the validated dataset for the scope of this study.

*Calculations.* Descriptive statistics were used to summarize the data. Continuous variables are presented as medians with interquartile ranges (IQR), and categorical variables as counts and percentages.

Empirical treatment was defined as the antibiotic initiated at the first visit before urine culture results were available. Across all datasets, a rule-based interpretation was applied:

- Guided treatment was defined as the antibiotic therapy that was modified or newly initiated when the diagnosis of febrile UTI was supported.
- Discontinuation was defined as stopping the empirical antibiotic when a UTI was considered unlikely.

The decision to continue with guided treatment or discontinue treatment, was based on clinicians’ assessment at follow-up, typically after urine culture result became available.

For validation purposes, prescribing outcomes in the Uppsala dataset were classified through clinician-adjudicated review of the electronic medical records and served as the reference standard for evaluation of the rule-based algorithm. This validation assessed the appropriateness of prescribing outcomes based on documented clinical reasoning, including alternative diagnoses, rather than solely the observed treatment decisions.

Two prescription indices were calculated to reflect the quality of antibiotic decision-making:

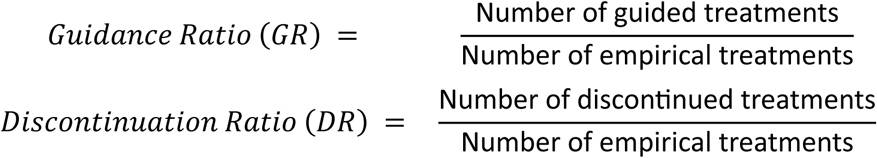

For the Uppsala dataset, ratios were calculated using the validated classification and for all study sites, guidance and discontinuation rates were calculated using rule-based algorithms. Further, the test performance of the rule-based approach was evaluated by calculating sensitivity and specificity in comparison with the validated classification. For this dataset only, monthly and overall ratios for the study period were derived from both the validated and the rule-based datasets. Agreement between the two approaches was assessed using the mean absolute error, bias, and standard error across overlapping time periods. Limits of agreement were defined as bias ± 1.96 SD according to the Bland–Altman method. [5] Lin’s concordance correlation coefficient (CCC) was calculated to assess overall concordance between the two GR measures. [6] To define the expected variation of the calculated indices, monthly values of the GR and DR were summarized using the 25th–75th percentile interval of the validation dataset to represent typical variation. The 10th–90th percentile interval was used to illustrate broader fluctuations over time. Values outside this range were interpreted as potential deviations from expected prescribing patterns rather than statistical outliers. (Figure 1)

**Figure 1.**
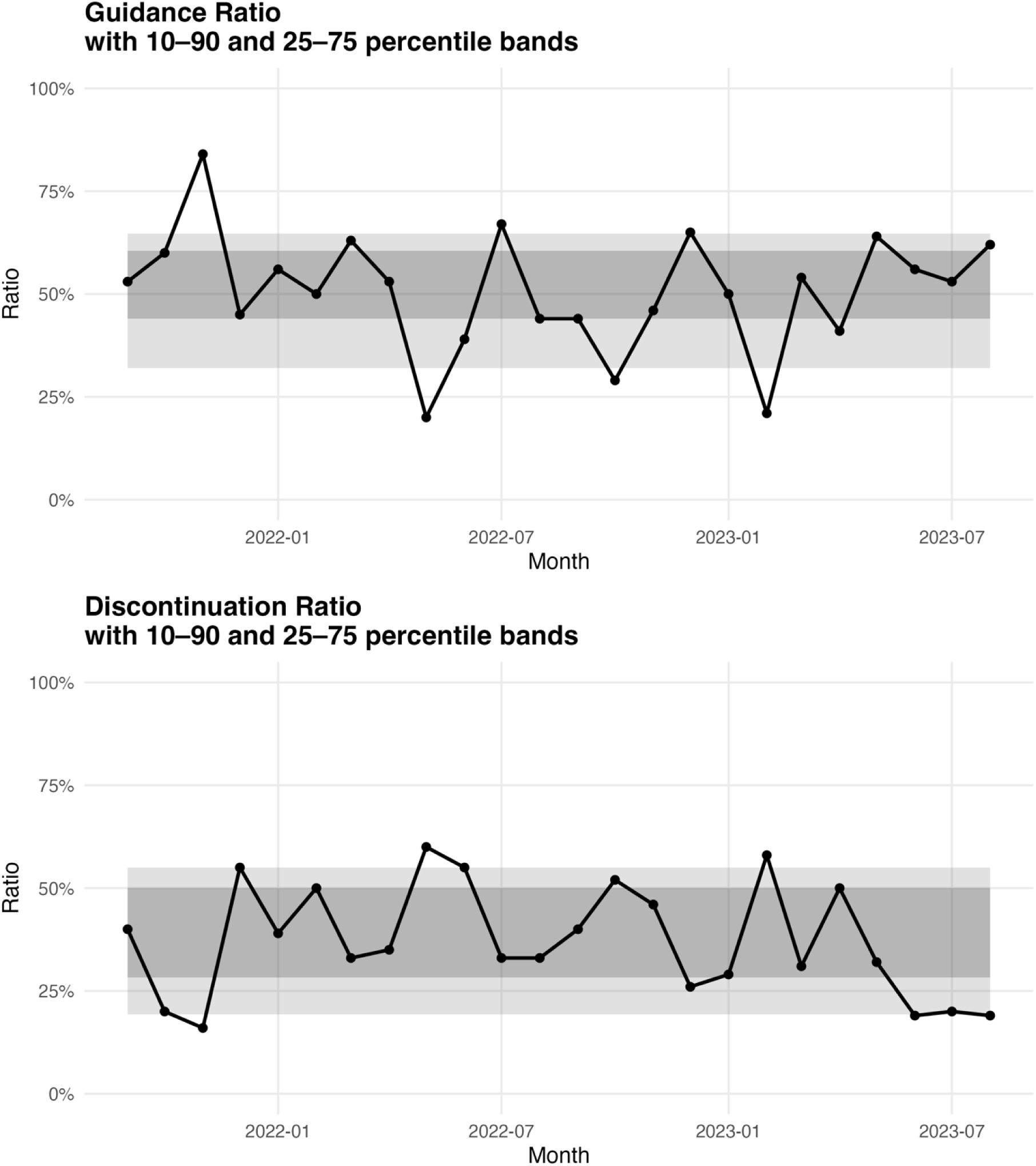
*Monthly trends in prescribing for rule-based ratios from Uppsala dataset*. Guidance Ratio (GR) and Discontinuation Ratio (DR), were calculated per month based on all empirical antibiotic treatments for suspected febrile urinary tract infection. Shaded areas represent percentile target ranges for each index: the dark grey band indicates the interquartile range (25th–75th percentile) and the light grey band the wider 10th–90th percentile range.

To explore whether prescribing decisions aligned with the strength of diagnostic evidence, an exploratory classification based on commonly used clinical and laboratory indicators was constructed. One point was assigned for each fulfilled criterion: body temperature > 38.5 °C, leukocyte esterase ≥ 2+, positive nitrite, C-reactive protein (CRP) > 70 mg/L, and significant growth of a relevant uropathogen in urine culture. [7] The total number of fulfilled criteria (0–5) reflected the overall level of diagnostic support for febrile UTI at the time of reevaluation. Missing values were treated conservatively as zero points. The score was used solely for exploratory stratification of the GR and applied across all study sites.

*Software and reproducibility*. Data were compiled and preprocessed in Microsoft Excel (Microsoft Corp., Redmond, WA, USA) and subsequently analyzed and visualized in R version 4.5.1 (R Foundation for Statistical Computing, Vienna, Austria).

## Results

### Study population from all sites

A total of 909 febrile UTI episodes were included Uppsala (n = 431), Solna (n = 336) and Huddinge (n = 142). The median age was 2 years (IQR 0.7–5), and 84% of patients were girls. Baseline clinical characteristics were similar across sites. Significant bacterial growth was found in 426 of 861 evaluable urine cultures (49%). (Table 1)

**Table 1.**
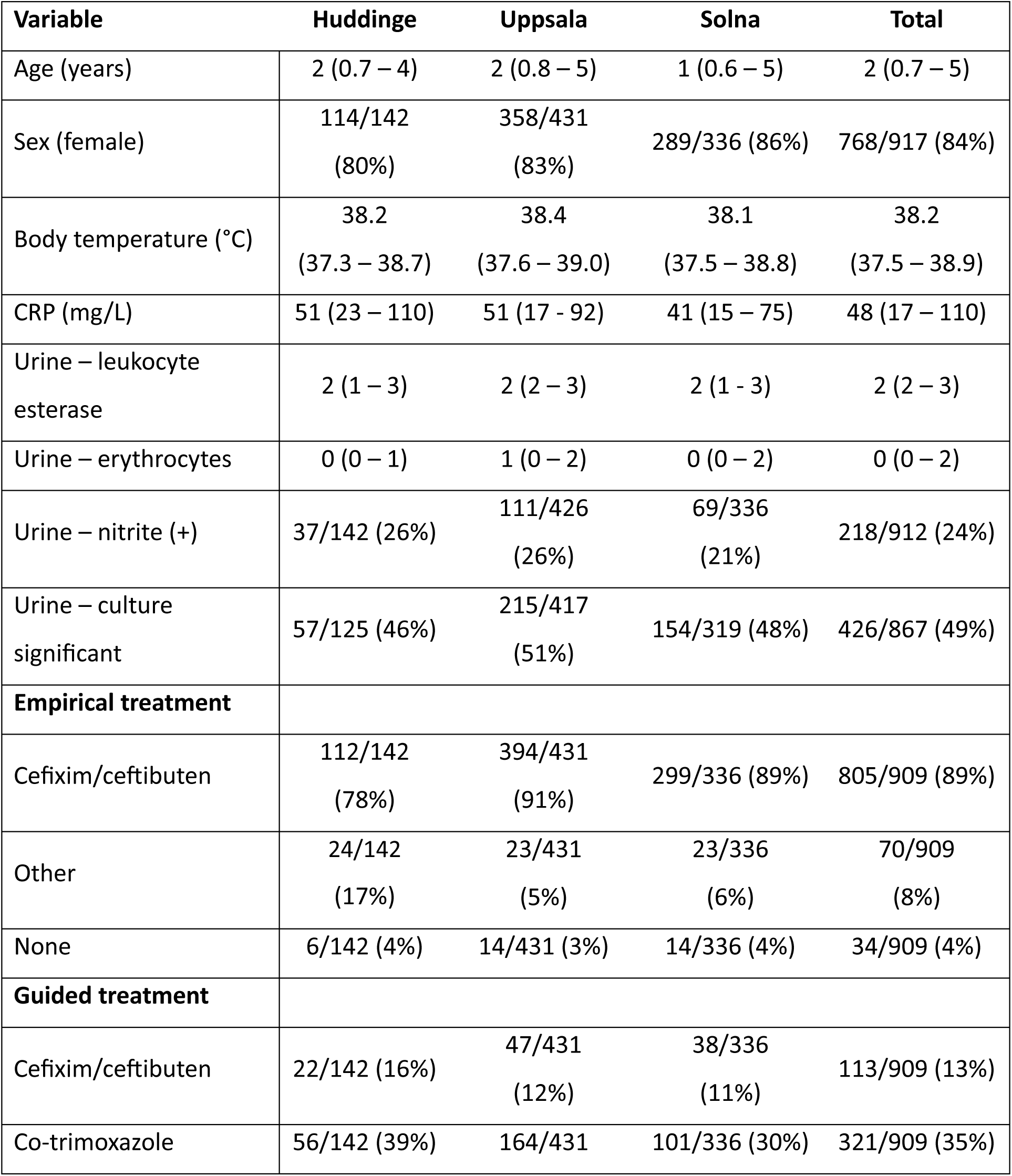
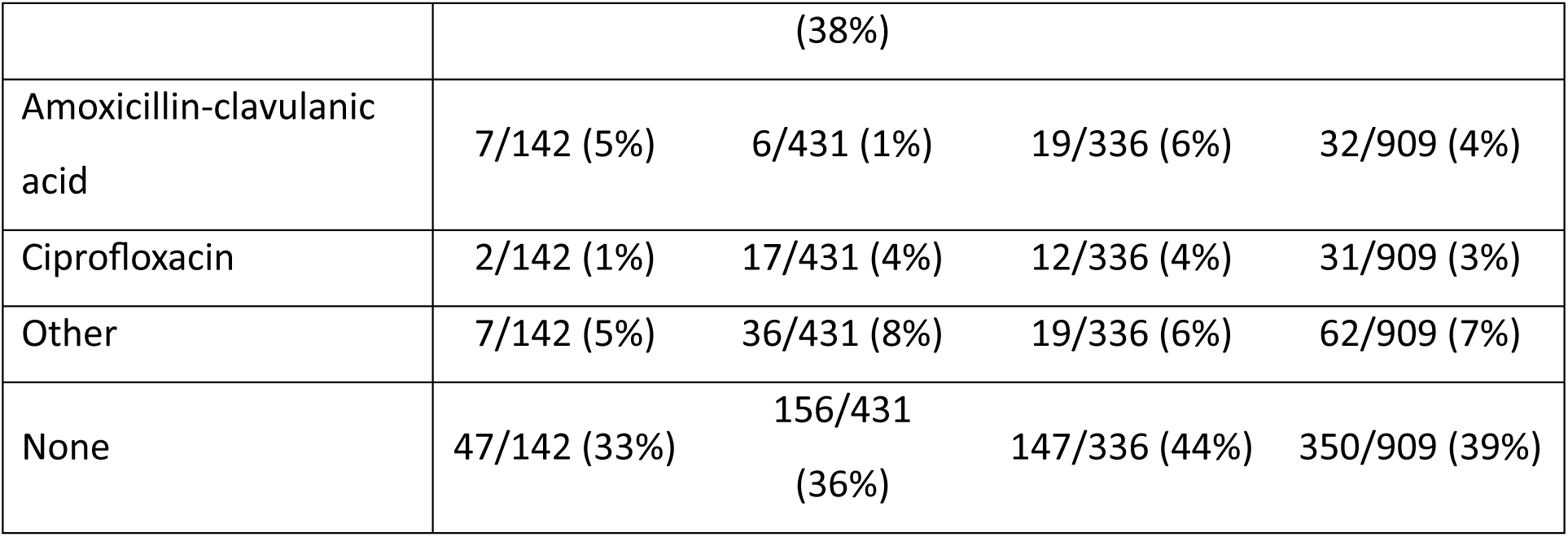
Baseline characteristics of the rule-based dataset for alla study sites. Data from pediatric patients with febrile urinary tract infection, collected from three university-affiliated emergency departments in Sweden. Continuous variables are presented as median and interquartile range (IQR), and categorical variables as counts and percentages.

Across all three study sites, cefixime or ceftibuten were the predominant empirical treatments, accounting for 805 of 909 episodes (89%). Other empirical agents were used infrequently (70/909, 8%), with the most common being co-trimoxazole, amoxicillin–clavulanic acid, and ciprofloxacin. No empirical antibiotic was initiated in 34/909 (4%) of cases. Among guided treatments, co-trimoxazole was the most frequently prescribed follow-up therapy (321/909, 35%), followed by cefixime/ceftibuten (113/909, 13%). Amoxicillin–clavulanic acid and ciprofloxacin were used in smaller proportions (32/909, 4% and 31/909, 3%, respectively), with other agents comprising 62/909 (7%). No guided antibiotics were prescribed in 350/909 (39%) of episodes.

### Prescribing outcomes and indices for the validation dataset

The Uppsala validation dataset included 431 empirically treated episodes of suspected febrile UTI. Of these 431 episodes, 273 (63%) were classified as guided based on urine culture results and clinical follow-up. Discontinuation occurred in 119 cases (28%), 39 episodes (9%) lacked sufficient follow-up documentation. The rule-based algorithm classified 219/431 (51%) episodes as guided, 156/431 (36%) as discontinued, and 56/431 (13%) as unchanged continuation. When compared with the validated classification, the rule-based approach demonstrated a sensitivity of 0.78 and a specificity of 1.00.

Monthly comparisons between the validated GR and the rule-based GR showed that the rule-based measure consistently followed the same overall pattern, but with lower absolute values. The validated GR ranged from 39% to 89%, whereas rule-based GR ranged from 20% to 84%. The largest discrepancies occurred in months with lower GR values (e.g., May 2022: 39% *versus* 20%; February 2023: 50% *versus* 21%). Agreement was closer in several months with higher GR values (e.g., November 2021: 89% versus 84%). Across the full study period, the mean absolute error between GR and rule-based GR was 0.20, with a negative bias indicating systematic underestimation by the rule-based algorithm. The limits of agreement were wide, and Lin’s concordance correlation coefficient (0.39) indicated weak concordance, although the rule-based GR captured the temporal pattern. To quantify the structural difference between the two measures, a simple linear regression with validated GR as the dependent variable was fitted. This yielded the calibration function: validated GR ≈ 0.349 + 0.699 × (rule-based GR). Applying this function to the rule-based values reduced the monthly discrepancies and produced corrected estimates that closely approximated the validated GR across the entire study period. (Figure 2)

**Figure 2.**
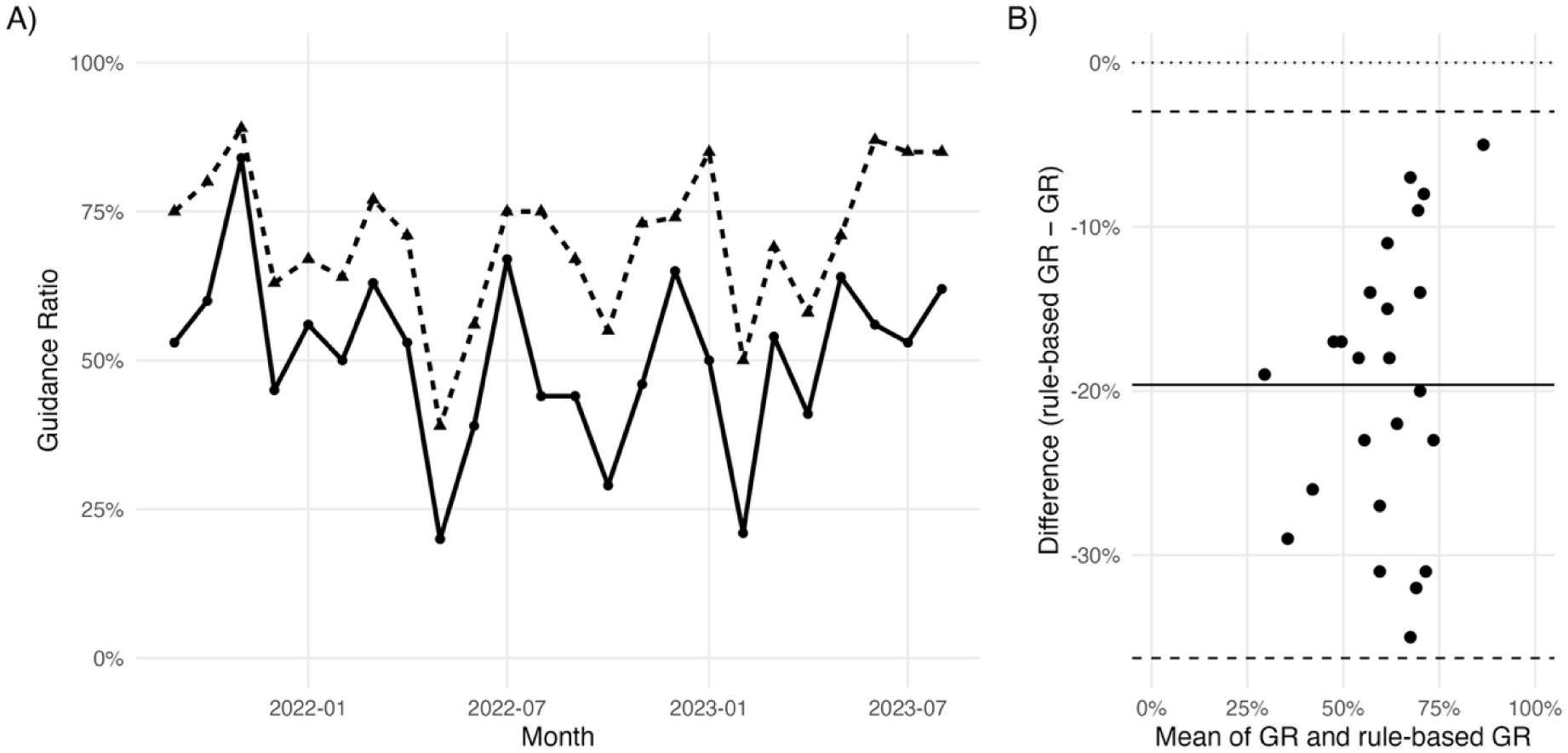
Comparison between Guidance Ratio (GR) based on validated dataset and rule-based proxy GR from Uppsala. (A) Monthly values of GR from validated dataset (solid line) and rule-based GR (dashed line) from September 2021 to August 2023. (B) Bland–Altman plot showing the difference between GR from validated dataset and rule-based GR (validated minus rule-based) plotted against their mean values. The solid line indicates the mean difference (bias), and dashed lines indicate the limits of agreement (±1.96 SD).

Compared to the monthly rule-based GR, DR showed the expected inverse pattern and ranged from 16% to 60%. Whereas most monthly estimates fell within the predefined 10th–90th percentile range, several showed marked deviations, particularly May 2022 (GR 20%, DR 60%) and February 2023 (GR 21%, DR 58%). Despite these fluctuations, no consistent upward or downward trend was observed, indicating overall stability in prescribing behavior over time. (Figure 1)

The overall rule-based GR was 75/142 (46%) in Huddinge and 153/336 (45%) in Solna, with a pooled multicenter value of 447/909 (49%). When stratified by the diagnostic evidence score, GR increased consistently with a higher number of fulfilled criteria across all sites. At intermediate diagnostic levels (e.g., two fulfilled criteria), Huddinge showed slightly higher GR values than Solna and Uppsala. The overall rule-based DR was 47/142 (33 %) in Huddinge and 147/336 (44 %) in Solna, with a pooled multicenter value of 350/909 (39 %). (Table 2)

**Table 2.** Rule-based Guidance Ratio (GR) by exploratory diagnostic evidence strata across study sites.

| Diagnostic criteria | Huddinge | Uppsala | Solna |
| --- | --- | --- | --- |
| 0 | 7/13 (54%) | 2/24 (8%) | 4/23 (17%) |
| 1 | 15/39 (38%) | 21/84 (25%) | 18/98 (18%) |
| 2 | 21/45 (47%) | 54/130 (42%) | 45/101 (45%) |
| 3 | 15/19 (79%) | 71/107 (66%) | 39/61 (64%) |
| 4 | 11/18 (61%) | 45/55 (82%) | 34/38 (89%) |
| 5 | 6/8 (75%) | 26/31 (84%) | 13/15 (87%) |
| Overall GR | 75/142 (46%) | 219/431 (51%) | 153/336 (45%) |

## Discussion

This multicenter validation study demonstrates that registry indicators can capture clinically meaningful patterns in antibiotic decisions after diagnostic reassessment in pediatric febrile UTI, while systematically underestimating concordance to guideline recommendations. By benchmarking measures based on registry data against reference outcomes established through clinical review, we show how calibration can transform routinely collected prescribing data into interpretable indicators for antimicrobial stewardship. This approach addresses a key gap in current stewardship surveillance, where most available metrics quantify antibiotic exposure rather than the quality of decisions made after diagnostic reassessment, which ultimately drive selection pressure and antimicrobial resistance at the population level.

Previous stewardship evaluations have highlighted the limitations of exposure-based metrics for assessing prescribing quality, as consumption measures cannot account for clinical context or appropriateness of therapy. [8–11] In addition, systematic reviews of stewardship indicators have shown that validated outcome measures remain rare and that testing against clinically meaningful outcomes is needed. [8,11] In line with these observations, our findings show that indicators reflecting diagnostic decision making after diagnostic reassessment capture clinically meaningful prescribing patterns that are not visible in measures based on prescribing volumes. Across the three study sites, most antibiotic episodes resulted in either guided treatment (45–51%) or treatment discontinuation (33–44%). The rule-based algorithm showed high specificity (1.00) and moderate sensitivity (0.78) for identifying guided therapy and captured temporal patterns in monthly GR values, although absolute levels were underestimated. Similar discrepancies between algorithm-based stewardship indicators and adjudicated outcomes after clinical reevaluation have been reported in previous validation studies using electronic health record data. [9,10,12] Agreement improved after calibration, indicating that systematic bias in registry-based indicators can be addressed using established methods for method comparison and measurement error. [13,14]

The stepwise increase in GR and DR with stronger diagnostic evidence supports the construct validity of the indicators, as prescribing behavior followed clinically expected patterns across diagnostic strata. (Table 2) Across all study sites, GR increased with the number of fulfilled clinical and microbiological criteria, supporting its use as an indicator of prescribing quality. Lower GR values at low diagnostic certainty were accompanied by higher DR values, which is consistent with appropriate discontinuation of empirical therapy when infection was considered unlikely. Similar associations between diagnostic certainty and antibiotic continuation have been described in pediatric febrile UTIs and other empirically treated infections, where post-discharge reassessment plays a key role in stewardship efforts. [3,4]

In the present study, both indices showed substantial month to month variation but no consistent upward or downward trend over time, suggesting overall stability in prescribing practice during the study period. (Figure 1) This temporal pattern indicates that indicators based on registry data can capture meaningful signals related to clinical decisions over time, even when absolute levels are imperfectly estimated. To capture aspects of clinical decision making not available in structured registry variables, more advanced approaches have been proposed. These include further development of electronic data structures and automated analysis of clinical free text using machine learning methods, and may reduce systematic misclassification in future implementations. [2,9,10]

Three months fell outside the percentile bands in Figure 1, providing potential signals for stewardship review. The available data did not allow a definitive explanation for these outliers, and we found no association with prescribing patterns or care seeking volume. UTI case volumes were comparable across periods, and the deviations were directionally consistent with microbiological findings, with more positive cultures during the high GR periods and fewer during months with low GR. We lacked data on laboratory reading practices and sampling quality and therefore cannot exclude that variation in specimen collection or interpretation contributed to these patterns. The deviations were limited to single months, which is reassuring, as sustained deviations over longer periods would be more concerning.

Previous studies have noted that antibiotic indicators that rely on registry data provide limited insight into the clinical reasoning behind continuation or discontinuation decisions. Discrepancies between rule-based classifications and clinician assessments are common, particularly when follow-up documentation is incomplete or delayed. [9,10] In line with these observations, the rule-based approach in our study captured overall prescribing patterns well but was less reliable for identifying treatment discontinuation in episodes without microbiological support, highlighting a limitation of relying solely on structured registry variables for outcome classification. At the same time, international stewardship initiatives, including the DRIVE AB programme, have shown that existing quality indicators predominantly target structure and process measures, while validated outcome indicators remain rare. [15] Many proposed indicators require clinical information to assess appropriateness, which is typically not available in prescribing registries, underscoring the need for validation studies that benchmark rule-based and registry-derived classifications against clinician documented outcomes.

A key strength of this study is the availability of detailed follow-up information in one of the participating regions, which allowed clinically verified outcomes to be used as a reference standard. Including all empirically treated febrile UTI episodes from several centers provides a broad and representative overview of prescribing patterns. In addition, the use of both aggregated and monthly indicators allows for a more nuanced interpretation of registry-based measures over time.

A limitation is that validated outcome classifications were only available for one region, and the results may therefore partly reflect local follow up and documentation practices. Registry-based data are limited in their ability to capture all aspects of clinical reasoning, which likely contributed to some of the observed discrepancies between rule-based and clinically verified outcomes. Documentation completeness varied, and approximately 10% of episodes lacked sufficient follow up information for full validation.

Rule-based registry indicators reflected overall prescribing patterns in pediatric febrile UTI but underestimated guidance after diagnostic reassessment when compared with clinical verified outcomes. Validation against clinical follow-up data demonstrated that GR and DR respond to diagnostic certainty, supporting their use as complementary stewardship measures for monitoring treatment decisions. The validation framework may be transferable to other empirically treated infections and health systems with linked prescribing and microbiology data.

## Data Availability

Data is available on contact with author upon reasonable request.

## Author contributions

The authors contributed to the study as follows: study conception and design: SS, AD, JL; data collection: AA, AD, KG, ES; calculations and visualization: SS, KG, AA. SS and KG took lead in writing the manuscript; all authors provided critical feedback and helped shape the research throughout all study phases and contributed significantly to the final manuscript.

## Funding

This study was supported from the ALF agreement between Uppsala University and Region Uppsala and a grant from the Swedish Research Council (2019-05909).

## Conflict of interests

The authors declare no conflict of interests.

## Availability of data and materials

The datasets used and/or analysed during the current study are available from the corresponding author on reasonable request.

## Notes

### Competing Interest Statement

The authors have declared no competing interest.

### Funding Statement

This study was funded by:
Uppsala Universitet, Sweden, ALF-1037235
Vetenskapsradet, 2019-05909

### Author Declarations

The study was approved by the Swedish Ethical Review Board (document ID 2020-03297).

